# Viral Dynamics Matter in COVID-19 Pneumonia: the success of early treatment with hydroxychloroquine and azithromycin in Lebanon

**DOI:** 10.1101/2020.05.28.20114835

**Authors:** Amanda Chamieh, Claude Afif, Gerard El-Hajj, Omar Zmerli, Isabelle Djaffar-Jureidini, Roy A. Raad, Raja Ashou, Georges Juvelekian, Jean-Marc Rolain, Eid Azar

**Affiliations:** Aix-Marseille Université, AP-HM, Institut de Recherche pour le Développement (IRD), Microbes Evolution Phylogeny and Infections (MEPHI), Institut Hospitalo-Universitaire Méditerranée-Infection, Marseille, France; Division of Infectious Diseases, Saint George Hospital University Medical Center, Beirut, Lebanon; Department of Radiology, Saint George Hospital University Medical Center, Beirut. Lebanon; Department of Laboratory Medicine, Saint George Hospital University Medical Center, Beirut, Lebanon; Department of Pulmonary and Critical Care Medicine, Saint George Hospital University Medical Center, Beirut, Lebanon

**Keywords:** COVID-19 pneumonia, Viral Load Dynamics in SARS-CoV-2, Hydroxychloroquine and Azithromycin in COVID-19

## Abstract

1.

**Background/Purpose:** We share our experience in COVID-19 pneumonia management at Saint George Hospital University Medical Center (SGHUMC) in Lebanon. In the absence of a standard of care, early diagnosis and opt-in therapy with Hydroxychloroquine and Azithromycin were offered.

**Methods:** We reviewed records of COVID-19 pneumonia patients from March 16-April 26 2020. Based on NEWS score, we stratified patients as A: low B: medium, and C: high clinical severity and obtained pharmacotherapy data. Chest-CT-severity-score (CTSS) was used. We defined clinical cure as resolution of symptoms and biomarkers and virologic cure as a PCR above 35 cycles(Ct).

**Results:** We recorded 21 COVID-19 pneumonia patients of whom 19 opted for treatment. Clinical symptoms and laboratory markers at presentation did not significantly correlate with severity. Lower initial viral load significantly correlated with lower levels of clinical and radiological severity (p=0.038). Virologic cure, Ct>35, by day 10, was only 33% in high severity significantly less than categories A and B. We observed 100% clinical cure at day 10 in Category-A, 67% in B, and 33% in C(p<0.05). Patients with the lowest severity had the fastest virologic cure in a mean of 5.8 days from diagnosis, shortest hospitalization and earlier radiological improvement(p<0.005). Ultimately, 18 patients were discharged home in good condition and one remains in the ICU.

**Conclusion:** Viral dynamics matter in COVID-19 pneumonia. An early control of replication may be crucial in averting complications. Early administration of Hydroxychloroquine and Azithromycin potentially explains our 94.7% success rate in treating a fairly complex cohort of COVID-19 pneumonia.

## 2. Introduction

SARS-CoV-2 is a novel coronavirus that is taking the world as we know it by storm.

In Lebanon, on February 21 2020, SARS-CoV-2 was first diagnosed in a female returning from the epicenter in Iran. Subsequently, the Lebanese government closed early on schools and enterprises, banned gatherings, and advised social distancing and home quarantine. Until April 26 2020, Lebanon is at 704 cases in a country of an estimated 6.7 million inhabitants.^1^

Globally, the approaches to managing COVID-19 pandemic are diverse. One method advocates conservative, home-stay for mild cases while reserving diagnostics and medical intervention for severe cases. Another main strategy is almost opposite and relies on early diagnosis by offering RT-PCR testing^2,3^.

At Saint George Hospital University Medical Center (SGHUMC) in Lebanon, we formed the SGHUMC COVID-19 response team and carefully reviewed existing evidence and management strategies. We opted for an early detection approach.

We tested all incoming patients with an acute respiratory, febrile illness or known exposure for SARS-CoV-2 by RT-PCR, traced their contacts, and performed prompt chest CT scan imaging as indicated.

In the absence of a standard of treatment for COVID-19, we assessed available therapies. Hydroxychloroquine and azithromycin are readily available promising combination with a familiar pharmacologic profile. On the other hand, lopinavir/ritonavir lacks significant clinical benefits and remdesivir use is restricted to clinical trials or compassionate use. These two options above are not available in Lebanon. IL-6 pathway inhibitors and convalescent plasma treatment are in trial reserved for advanced COVID-19. Therefore, we agreed to consider them on a case-by-case basis.^4–9^

In summary, patients with COVID-19 pneumonia chose either no pharmacological treatment or the combination of Hydroxychloroquine and azithromycin. Patients opting for the Hydroxychloroquine and azithromycin regimen consented verbally after having the risks and potential benefits of this regimen well explained to them.

In this retrospective study, we share our experience with COVID-19 pneumonia clinical and radiological presentations and outcomes, course of illness, and viral dynamics.

## 3. Methods

### 3.1. Study setting and Design

We conducted this retrospective study at SGHUMC in Beirut, Lebanon, from March 16, 2020 until April 26, 2020.

Patients were received via our Emergency Room and Corona Clinic or as transferals requiring urgent critical care, some of which were COVID-19 confirmed.

We obtained exposure and contact history of all incoming patients with respiratory or febrile illnesses and performed a physical exam. Nasopharyngeal and oropharyngeal swabs were obtained for SARS-CoV-2 RT-qPCR. Chest CT imaging was done within 3 days of diagnosis.

The SGHUMC response team was composed of infectious disease physicians, pulmonary and critical care physicians, radiologists, infection preventionists, nursing department, laboratory department, and a representative from hospital leadership.

Clinical and virologic cure was assessed at day 10 from diagnosis. Patients were followed until complete clinical and virologic clearance, defined as the overall outcome.

### 3.2. Inclusion Criteria

In our analysis, we included all patients that were positive for SARS-CoV-2 by RT-qPCR with findings of pneumonia on chest CT scan and had received Hydroxychloroquine and azithromycin for at least 72 hours.

### 3.3. Data Collection

We retrieved data from the electronic database of the COVID-19 clinic and emergency care service. We recorded patient age, gender, history of cardiovascular disease, hypertension, chronic lung conditions, diabetes mellitus, malignancy, and any immunosuppressive therapy. We also recorded possible exposure history and reported symptoms of cough, fever, headache, shortness of breath, chills, myalgias, sore throat, rhinorrhea, and diarrhea.

The time from symptom onset to diagnosis, imaging and treatment initiation, length of hospital stays, the need for oxygenation, and whether the patient was transferred to the intensive care unit (ICU) were retrieved. Laboratory markers included CBC, Creatinine, CRP, Troponin, Procalcitonin, Urea, Na, K, Ca, Mg, AST, ALT, Bilirubin, LDH, D-Dimer, Ferritin, and Fibrinogen depending on the case.

We considered day zero as the day of diagnosis of COVID-19 by RT-qPCR. At day 10, patients were assessed for clinical and virologic cure. Some patients underwent a repeat chest CT scan.

### 3.4. Clinical Assessment

We used the National Early Warning Score (NEWS) and chest CT scan findings to assess acute clinical severity on admission. ^10^ The NEWS is based on physiologic parameters that include respiratory and heart rate, blood pressure, oxygen saturation, use of supplemental oxygen, and state of consciousness. A NEWS score of 0-4, 5-6, and 7-15 respectively corresponded to low, medium, and high scores. For the sake of data analysis, we then divided our cohort into categories A, B, and C according to low, medium, and high NEWS score.

We define clinical cure as the resolution of symptoms, clinical stabilization, and normalization of inflammatory markers.

Patients were managed as inpatients or followed as outpatients depending on their clinical severity. Outpatients were discharged with strict instructions and education, and were called for follow up every 2 days.

### 3.5. PCR Assay

RNA was extracted from nasopharyngeal/oropharyngeal 158 samples. Real-time reverse transcriptase PCR (RT-qPCR) detection of SARS-CoV-2 was done using the LightMix® Modular SARS-CoV-2 (COVID19) E, N, and RdRP-genes (Tib Molbiol, Berlin, Germany) as previously described.^11,12^ We used the cycle number threshold values (Ct) as an indicator of the number of copies of SARS-CoV-2 present in our samples. We define virologic cure as 2 consecutive SARS-CoV-2 RT-qPCR with Ct>35, as previously described.^13^

We considered a patient with virologic cure by day 10 of diagnosis a fast responder and a slow responder if more than 10 days. All patients were followed with serial RT-PCR till negative, beyond limit of detection of the test, as part of SGHUMC infection control strategy.

We also tested for other pathogens using Biofire® FilmArray® Respiratoy 2 Panel (BioMerieux, France) following manufacturer’s instructions.

### 3.6. Radiologic Assessment

Non-enhanced low-dose chest CT scans were obtained in supine position on a "GE Healthcare: Discovery CT750 HD” CT scanner, dedicated for confirmed or suspected COVID-19 patients. Patients were instructed to hold their breath, and acquisition was made at end-inspiration.

COVID-19 typical findings were recorded, including: (a) ground-glass opacities, (b) dense airspace consolidation, (c) interlobular septal thickening, (d) vascular enlargement, (e) peripheral, subpleural distribution, and (f) multifocal/multilobar/bilateral involvement. The presence of pleural effusion and other findings atypical for pneumonia was also assessed. Chest CT severity score (CTSS) was determined to estimate the severity of lung involvement in patients with COVID-19 pneumonia. The scoring system was adapted from similar scoring systems used in recent COVID-19 literature. ^14,15^ A score of 0, 1, 2, or 3 (corresponding to involvement of 0%, 0-25%, 25-50%, and >50% respectively) was assigned for each lobe of the 5 lung lobes with a maximum total score of 15. Therefore, if a patient has a CTSS of 15, it indicates that there is more than 50% involvement of the lung. Interval improvement and resolution is a decrease in CTSS and/or subjective decrease in ground-glass opacities density.

All images were anonymized then analyzed by two experienced radiologists (R.A. and R.R.) independently. In case of discrepant CT findings or CTSS, a final decision was reached by consensus.

### 3.7. Treatment

#### 3.7.1. Pharmacologic therapy for patients receiving Hydroxychloroquine and azithromycin

We verified for any drug-drug interactions, G6PD deficiency and contraindications, and we obtained an ECG. If QTc<460msec, treatment was initiated. ^16^ When QTcB>460msec, we consulted a cardiologist that ordered continuous cardiac monitoring by telemetry if needed. ECG was repeated on day 2 and day 10 upon completion of therapy.

We administered 600mg of Hydroxychloroquine per os per day for 10 days and a 500mg loading dose of azithromycin followed by 250mg/day for 4 days intravenously or orally.

#### 3.7.2. Non-pharmacologic Treatment

Supportive and symptomatic therapy, such as oxygen, was assessed individually.

### 3.8. Statistics

The Shapiro-Wilk test was used to assess normality. Data were expressed as mean ± SD and/or median [minimum; maximum]. The nonparametric Man-Whitney U or Kruskal-Wallis H test was applied to assess for differences between categories. The Wilcoxon signed-rank test was used to assess improvement in CTSS values on follow-up CT. A Spearman's rank-order correlation was used for interrater reliability between the radiologists' CTSS. The rank-based Jonckheere-Terpstra test was applied to trend the 3 clinical severity groups and cycle threshold numbers. P-values<0.050 were significant.

### 3.9. Ethical Statement

We obtained SGHUMC IRB approval for a retrospective review of the charts of COVID-19 patients. It is standard procedure that all patients sign a general informed consent for receiving medical acts and care.

## 4. Results

Out of 860 patients that presented to our COVID-19 clinic, 27 patients had SARS-CoV-2 RT-PCR positive nasopharyngeal swabs at our institution. Six of the 27 patients were asymptomatic with a normal chest CT scan. Two of the remaining 21 patients with COVID-19 typical pneumonia features on chest CT scan refused pharmacologic treatment and followed-up with their physicians. These 8 patients were excluded.

Finally, 19 patients were included, all opting for treatment with the combination of Hydroxychloroquine and azithromycin.

To note, two patients, not diagnosed nor initially treated at SGHUMC, were transferred to SGHUMC for critical care. Both had advanced COVID-19 and died within 72 hours of admission. These two patients were excluded.

The mean age in our study cohort was 57.8±16.4 years. Category A of low clinical severity was composed of 4 males and 6 females, with a median age of 51[28;85]. Category B was composed of 5 males and 1 female, median age 59[52;67]. Category C was composed of 2 males and 1 female, median 81[55;87]. No statistical difference observed in ages across the 3 categories. We observed a higher ratio of 11 males to 8 females.

As for comorbidities, 4 patients in category A had hypertension (HTN) and another 4 had dyslipidemia. In category B, one patient was a smoker with mild chronic bronchitis, and another one had coronary artery disease (CAD), HTN, and dyslipidemia. In category C, one patient had CAD, HTN and dyslipidemia while another was a smoker. None of the patients were known diabetics.

Only 1 patient with COVID-19 pneumonia of medium NEWS score was co-infected with RSV.

### 4.1. Clinical Course and Cure

The median duration from initial symptom onset to diagnosis of COVID-19 was 3.5[1;18] days in category A, 8[2;12] days in category B and 4[2;8] days in category C. This was not statistically significant.

At initial evaluation, there was no significant variation between symptoms reported by patients. They reported fever in 15/19, cough in 13/19, shortness of breath in 9/19, myalgias in 5/19, sore throat in 4/19, chills in 2/19, headache in 2/19, and 1/19 with fatigue. Anosmia was reported by 1 patient.

We also found no significant difference in laboratory markers among patients of all categories upon presentation.

#### 4.1.1. Clinical Course

The length of stay of patients in category A was a mean of 3±4.8 days, median 1[0;12] days, significantly lower than both category B at a mean of 11±4.8 days; median 12.5[3,16] days (p=0.032) and category C at a mean of 20±8.5 days, median of 19[12,29] days(p=0.015). No statistical significance in LOS between medium and high clinical severity.

There was no distinct pattern for progression of laboratory markers. However, when peak values are reached, it took 4-6 days to return to baseline. This was independent of initial clinical severity. Table 1 represents the results of available blood tests during the study period per category. On average, category A of low clinical severity had an overall significantly lower CRP, Procalcitonin, LDH, Ferritin, D-Dimer, and lymphopenia.

Table 1: *Laboratory Markers of Patients in Category A, B, and C of low, medium and high clinical severity*

#### 4.1.2. Clinical Cure at day 10

All patients in category A achieved clinical cure on day 10 compared to 4/6 in category B. In category C, only 1 patient achieved clinical cure at day 10. Thus, 100% clinical cure at day 10 in low clinical severity, 67% in medium severity, and 33% in high severity COVID-19. This trend was statistically significant (p=0.031).

#### 4.1.3. Overall Clinical Outcome

Eighteen out of 19 patients were discharged home in good condition, a clinical success rate of 94.7%. Only 1 patient in category C was still on mechanical ventilation. This patient is the eldest in the cohort at 87 years and had the highest NEWS score of 9 on admission.

None of the patients developed any drug related adverse events.

### 4.2. Virologic Load Dynamics

#### 4.2.1. Initial Viral Load

A higher initial SARS-CoV-2 RT-PCR Ct reflects a lower viral load. There was a statistically significant trend of higher Ct value with lower levels of clinical severity (p=0.038). Median Ct in category A was 33.5(13,38) which is significantly lower than that of category B at Ct= 30[23,35]. This was also significantly lower than the initial Ct in category C of 24[13,29] (Figure 1A).

**Figure 1:**
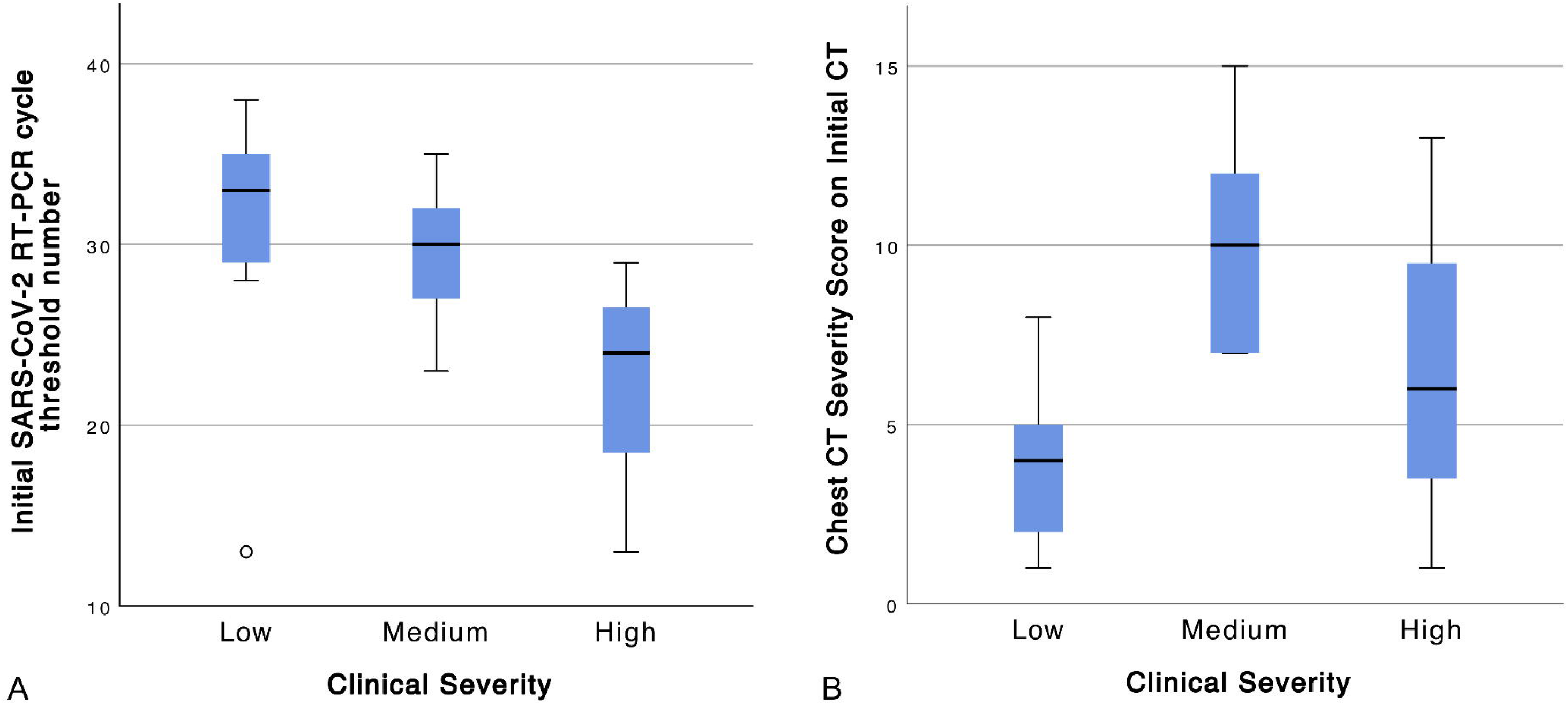
(A) Initial viral load: A higher initial SARS-CoV-2 RT-PCR cycle threshold number (Ct) reflects a lower viral load. There was a statistically significant trend of higher Ct value with lower levels of clinical severity (p=0.038). (B) Initial Chest CT Severity Score (CTSS): On initial CT, there was a statistically significant difference in CTSS between the 3 categories (p = 0.012). Patients of category B of medium clinical severity had higher CTSS than category A (low clinical severity) (p = 0.003) and category C (high clinical severity) (p = 0.192).

#### 4.2.2. Virologic Outcome at Day 10

In category A, PCR results were available for only 8/10 patients, and 6/8 patients achieved virologic cure at day 10 in a mean of 5.8±4.2 days. In category B, PCR results were available for 5/6 patients, and 4 achieved virologic cure in a mean of 9.2±4.2 days. In category C, only 1/3 patients achieved virologic cure in 4 days. (Figure 2)

**Figure 2:**
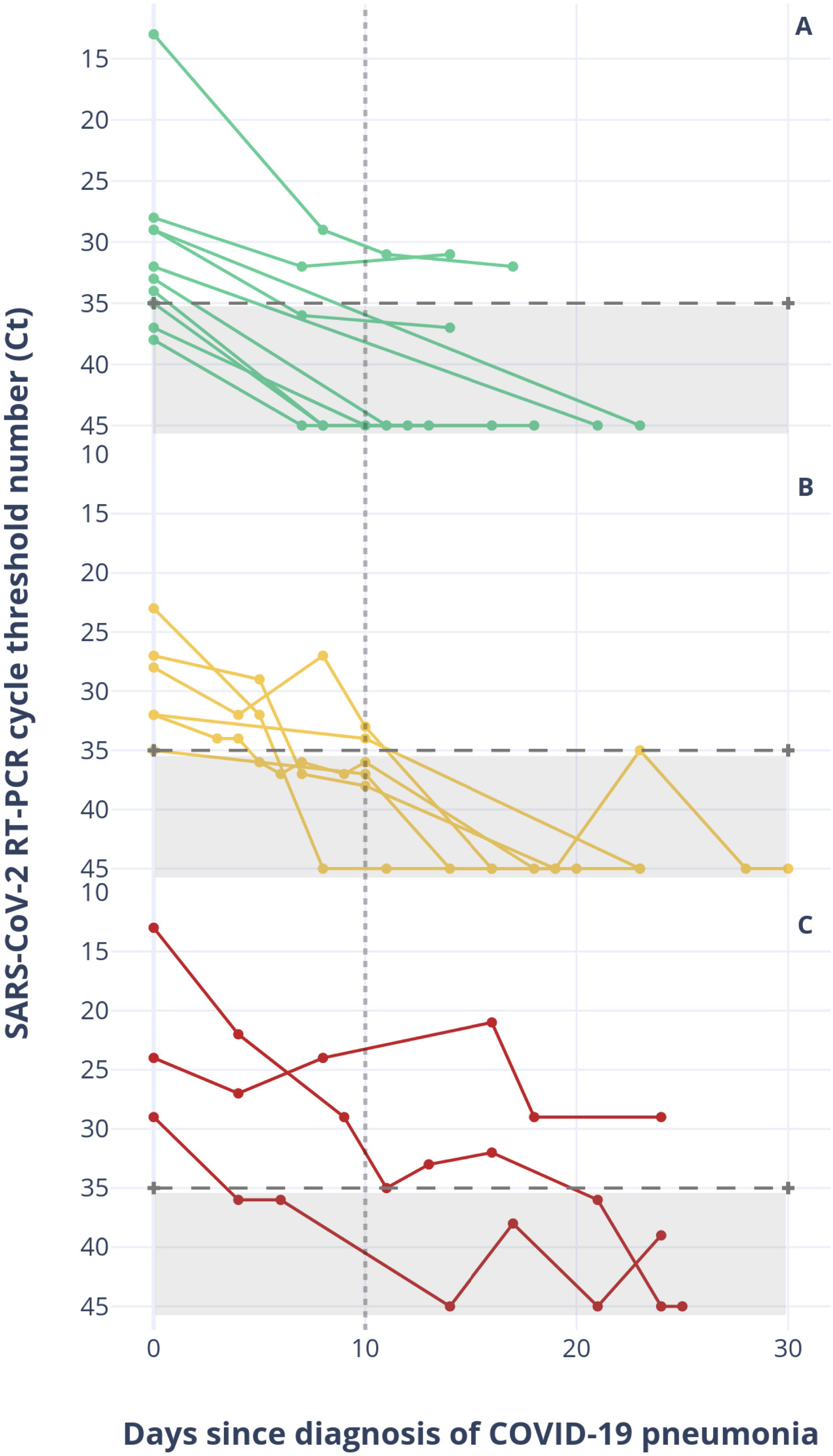
Viral Dynamics from day of diagnosis till negativity. *x-axis:* Days since diagnosis of COVID-19 pneumonia. *y-axis:* SARS-CoV-2 RT-PCR cycle threshold number (Ct) follow up according to low(A), medium(B), and high(C) clinical severity. Patients with lower clinical severity reached virologic cure significantly faster than those of higher severity (p=0.042).

#### 4.2.3. Overall Virologic Cure

At the end of the study, SARS-CoV-2 RT-PCR results were available for all patients. In category A, eight of ten patients achieved virologic cure, beyond the limit of detection of our test at mean of 14.8±7.7 days. In category B, all patients achieved virologic cure beyond limit of detection of our test and clinical cure at a mean of 16±5 days. As for category C, only 2 out of the 3 patients achieved virologic cure beyond limit of detection of our test on days 21 and 24. (Figure 2)

Patients with lower clinical severity reached virologic cure significantly faster than those of higher severity (p=0.042).

### 4.3. Radiologic Findings

There was a strong, positive correlation between the two readers R.A. and R.R., which was statistically significant (rs(36) = 0.958, p < 0.001).

All patients had a chest CT scan done within 3 days of COVID-19 diagnosis. To note, the median time from symptom onset to imaging was 3 days for categories A and C and 8 days for category B. (p = 0.842).

On initial CT, there was a statistically significant difference in chest CT Severity Score(CTSS) between the 3 categories (p = 0.012) (Figure 1B). Patients of category B showed the highest median CTSS of 10/15 compared to A and C (4/15 (p = 0.003) and 6/15 (p = 0.192), respectively). (Figures 3,4).

**Figure 3:**
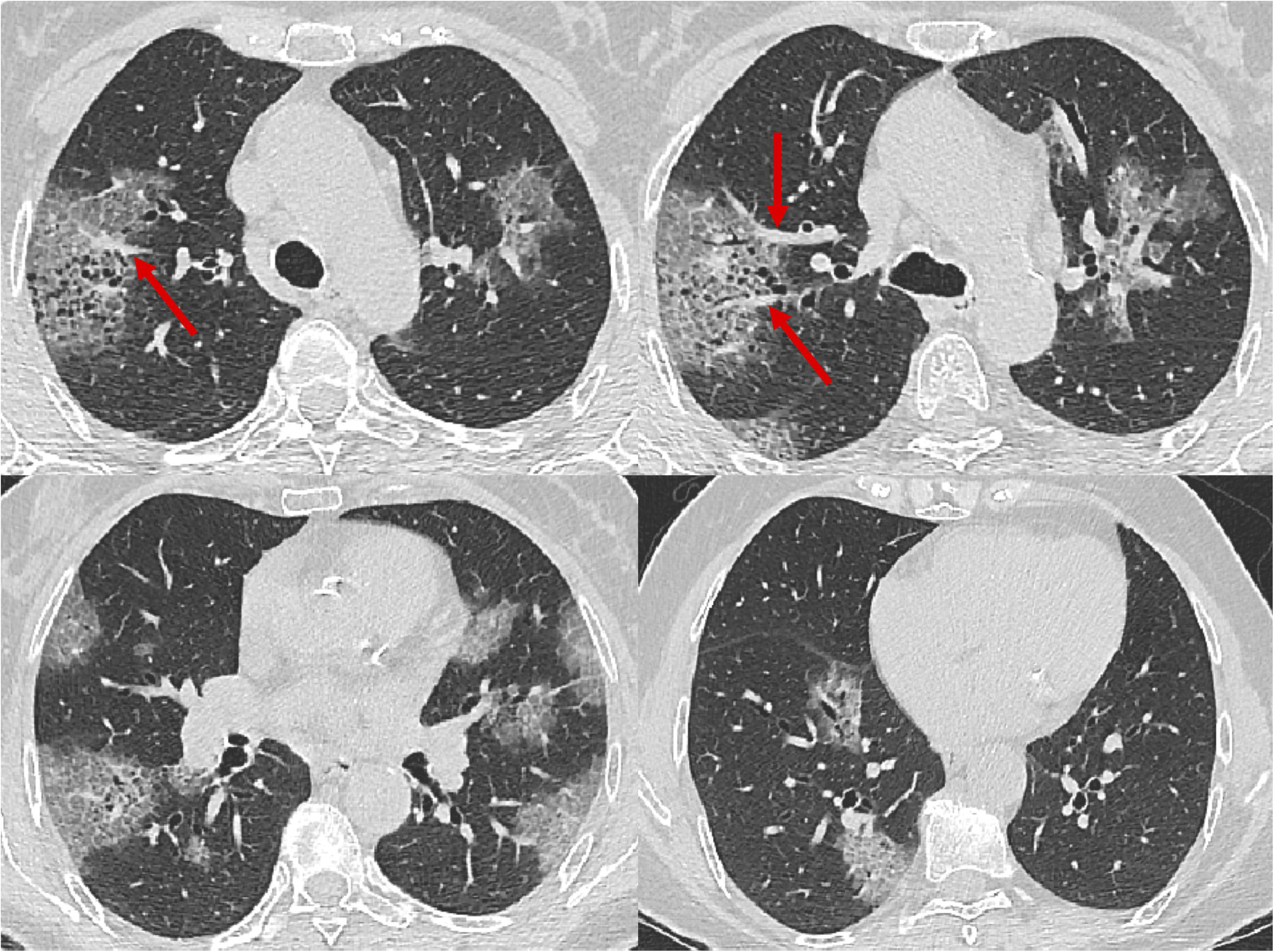
Axial serial chest CT images of a 66-year-old female with cough, fever, and shortness of breath (category A). Baseline chest CT demonstrating multilobar ground-glass opacities and dense consolidations predominantly in a peripheral subpleural distribution, with associated interlobular septal thickening (“crazy paving pattern”) and vascular enlargement sign (arrows). Chest CT Severity Score was estimated at 9/15. Clinically cured at day 10, and virologic cure confirmed after 21 days.

**Figure 4:**
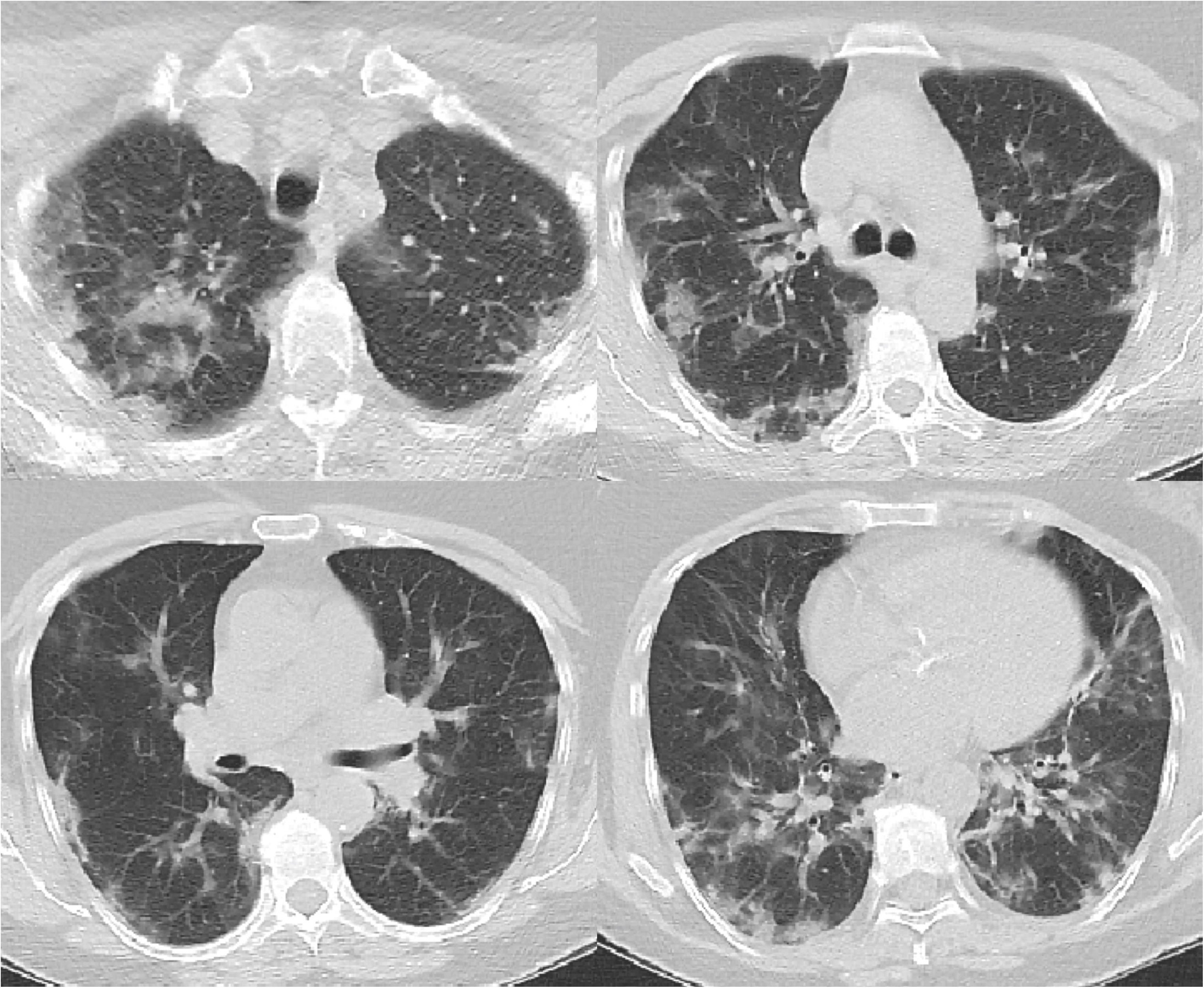
Axial serial chest CT images of a 59-year-old female (category B) with cough, fever, shortness of breath. Baseline chest CT demonstrating multilobar ground-glass opacities and dense consolidations predominantly in a peripheral subpleural distribution, with associated mild septal thickening. Chest CT Severity Score was estimated at 12/15.

Only 15 patients underwent follow-up chest CT scans at a median of 14 days (range 10-26 days). Thirteen (87%) patients showed significant interval improvement of which two patients showed complete resolution (p = 0.031). (Figure 5)

**Figure 5:**
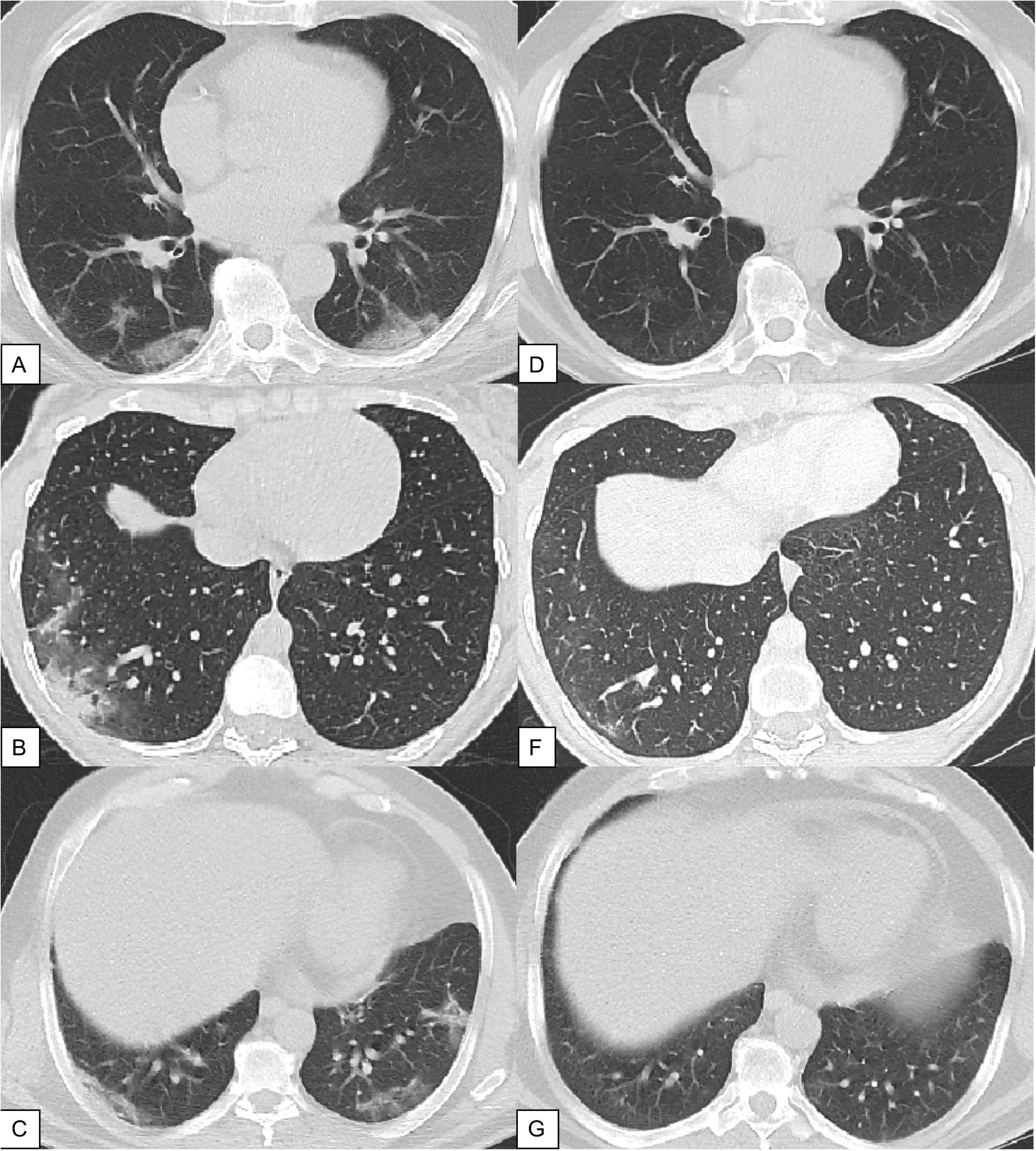
Three patients with COVID-19 pneumonia findings including peripheral subpleural ground glass opacities and dense consolidations in the lower lobes (A and C) and in the right lower lobe (B), with near-complete resolution on follow-up CTs (D,F and G). Chest CT Severity Score also improved on follow-up exams.

The remaining two patients with worsening features on second CT showed significant improvement on the subsequent third CT scan (Figure 6), obtained on day 20 and 29 respectively.

**Figure 6:**
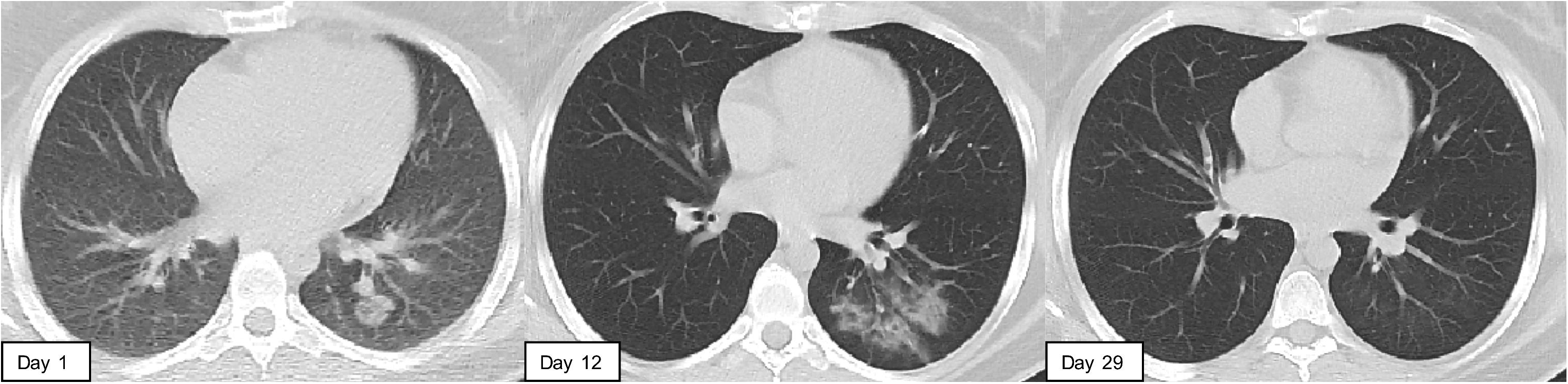
Chest CT of a 29-year-old female with fever, myalgia, and possible contact with COVID-19 positive case. Baseline chest CT (Day 1) showing a solitary rounded mixed attenuation opacity in the right lower lobe. Findings worsened on the initial follow-up chest CT (Day 12), with multiple new ground-glass opacities in the right lower lobe (B), then completely resolved on the subsequent follow-up CT (Day 29).

The median follow-up interval from time of diagnosis to radiological improvement for the 15 patients was: 14 (10-24), 12 (10-29), and 20 (14-26) days for categories A, B, and C respectively.

## 5. Discussion

At SGHUMC, 78% of patients diagnosed with COVID-19 had pneumonia on chest CT scan. Symptoms such as shortness of breath and dyspnea were not reliably predictive of disease severity. Our low threshold to perform chest CT scan was crucial in early diagnosis.

In our cohort, an initially low viral load was associated with less severe clinical and radiological findings, and faster viral clearance. All patients in category A achieved clinical cure in 10 days and had the fastest virologic cure in a mean 5.8 days from diagnosis. In contrast, only 56% of patients of higher clinical severity achieved clinical cure in 10 days, had longer hospital stays, and a more prolonged viral shedding. A recent study on 1061 patients with COVID-19 at IHU-Mediterranee Infection in Marseille, France showed that patients with higher initial viral load were less likely to have a low clinical severity and more likely to have poor virologic outcome.^17^Our results are consistent with their findings confirming that initial viral load and kinetics of viral clearance matter in COVID-19.

In many viral infections, early control of viral replication is crucial for better outcomes. Hydroxychloroquine has an antiviral effect that is proven in-vitro and in-vivo.^18–23^ We had a 94.7% success rate in treating a fairly complex cohort of COVID-19 pneumonia, with 18/19 patients discharged at baseline conditions. None of our 19 pneumonia patients had a cytokine storm, and all had a rapid decline within 4 days from peak inflammatory laboratory markers. The latter is likely a result of virologic control and possible early attenuation of cytokine storm. ^7,24,25^

Chest CT scan is a helpful diagnostic tool but also serves to monitor progression. Thirteen of 15 follow-up chest CT scans improved within 14 days. The remaining two showed improvement in a third scan beyond 20 days. In fact, all patients improved, showing either complete resolution or marked improvement in the CT findings, mainly ground-glass opacities and parenchymal consolidations. A follow-up chest CT may be necessary inpatients with COVID-19 to monitor for secondary organizing pneumonia and subsequent fibrosis, similar to outcomes seen with SARS and MERS.^14^ We will continue to monitor our patients for such complications despite the current significant radiologic improvement.

Our observational retrospective cohort is not designed to measure the direct impact of hydroxychloroquine and azithromycin treatment on the outcome of COVID-19 pneumonia. However, in 10 days, 80% of patients with varying severity of pneumonia reached clinical cure, and 70% achieved a virologic cure. Hence, early diagnosis of COVID-19 pneumonia coupled with administering Hydroxychloroquine and azithromycin treatment contributed to faster clinical and radiologic resolution and virologic clearance, potentially averting complications.

In conclusion, it is clear that higher viral load and prolonged viral shedding results in a longer and more complex clinical course. Our experience in early patient tracing, testing, chest imaging, and treatment of COVID-19 of mild, moderate, and severe pneumonia with Hydroxychloroquine and azithromycin resulted in a 95% success rate with no mortality. Further studies outside of a pandemic setting are needed to fully and fairly demonstrate the impact of early treatment of COVID-19 pneumonia with Hydroxychloroquine and azithromycin on overall patient outcome, long-term lung damage and reduction of the transmission of SARS-CoV-2.

## Data Availability

The data that support the findings of this study are available from the corresponding author upon reasonable request.

## Acknowledgements

This work was supported by the Saint George Hospital University Medical Center Beirut, Lebanon.

